# Direct and indirect effectiveness of mRNA vaccination against SARS-CoV-2 infection in long-term care facilities in Spain

**DOI:** 10.1101/2021.04.08.21255055

**Authors:** Susana Monge, Carmen Olmedo, Belén Alejos, María Fé Lapeña, María José Sierra, Aurora Limia, COVID-19 registries study group

## Abstract

**Objectives:** To estimate indirect and total (direct plus indirect) effects of COVID-19 vaccination in residents in long-term care facilities (LTCF).

**Design:** Registries-based cohort study including all residents in LTCF ≥65 years offered vaccination between 27 December 2020 and 10 March 2021. Risk of SARS-CoV-2 infection following vaccination was compared with the risk in the same individuals in a period before vaccination. Risk in non-vaccinated was also compared to a period before the vaccination programme to estimate indirect protection. Standardized cumulative risk was computed adjusted by previous documented infection (before the start of follow-up) and daily-varying SARS-CoV-2 incidence and reproductive number.

**Participants:** 573,533 records of 299,209 individuals in the National vaccination registry were selected; 99.0% had ≥1 vaccine-dose, 99.8% was Pfizer/BioNTech (BNT162b2). Residents mean age was 85.9, 70.9% were females. A previous SARS-CoV-2 infection was found in around 25% and 13% of participants, respectively, at the time of vaccine offer and in the reference period.

**Main outcome measures:** Documented SARS-CoV-2 infection identified in the National COVID-19 laboratory registry.

**Results:** Total VE was 57.2% (95% Confidence Interval: 56.1%-58.3%), and was highest ≥28 days after the first vaccine-dose (*proxy* of ≥7 days after the second dose) and for individuals naïve to SARS-CoV-2 [81.8% (81.0%-82.7%)] compared to those with previous infection [56.8% (47.1%-67.7%)]. Vaccination prevented up to 9.6 (9.3-9.9) cases per 10.000 vaccinated per day; 11.6 (11.3-11.9) if naïve vs. 0.8 (0.5-1.0) if previous infection. Indirect protection in the non-vaccinated could only be estimated for naïve individuals, at 81.4% (73.3%-90.3%) and up to 12.8 (9.4-16.2) infections prevented per 10.000 indirectly protected per day.

**Conclusions:** Our results confirm the effectiveness of mRNA vaccination in institutionalized elderly population, endorse the policy of universal vaccination in this setting, including in people with previous infection, and suggest that even non-vaccinated individuals benefit from indirect protection.

**Key messages:**

- COIVD-19 vaccination reduced the risk of documented SARS-CoV-2 infection in institutionalized elderly by 57.2% (56.1% to 58.3%), which increased to 81.2% (80.2% to 82%) for the fully vaccinated.
- In individuals naïve to SARS-CoV-2 vaccination reduced the risk by up to 81.8% and averted up to 11.6 cases per 10,000 vaccinated persons per day.
- Those with previous infection also benefited from a risk reduction of 57%, which translated in less than 1 infection averted per 10,000 vaccinated persons per day.
- Non-vaccinated individuals living in facilities where the majority (residents and staff) had been vaccinated showed a risk reduction similar to those actually vaccinated.

## Introduction

Since the beginning of the COVID-19 pandemic up to March 7 2021, 18,927 residents in long-term care facilities (LTCF) have died in Spain with confirmed COVID-19, and an additional 10,492 have died with compatible symptoms [1]. This means a cumulative mortality rate of 67 per 1,000 residents, accounting only for confirmed infections. This high vulnerability is due to the higher risk of exposure in dependents living in a closed institution but also to the higher severity of infection due to advanced age and presence of comorbidities. Indeed, one on every 5 cases of SARS-CoV-2 infection died in this setting [1].

COVID-19 vaccination in Spain started on December 27 with the Pfizer/BioNTech (BNT162b2) vaccine, for which LTCF -both residents and workers-were the first priority group [2]. The vaccination campaign coincided with the third COVID-19 epidemic wave, with national 14-day cumulative incidence increasing from less than 250 cases per 100,000 population by the end of 2020 to more than 1,000 by the end of January 2021 [3]. Vaccination started in facilities considered at higher risk, such as those that had never experienced a COVID-19 outbreak, had higher number of residents or more difficulties for implementing prevention and control measures. Vaccination teams visited the facilities and vaccination was universal, including those with previous SARS-CoV-2 infection. Vaccination was only deferred in people with active infection and, inconsistently, in people under quarantine. Acceptance has been very high, with 97.8% of all institutionalized persons (any institution type) having received at least one vaccine dose, and 88.8% two doses [4].

The Pfizer/BioNTech vaccine has shown an efficacy of 95% in preventing Covid-19 in randomized clinical trials [5]. However, elderly persons in general, and those institutionalized in particular, are not represented in randomized studies [6]. Therefore there is great interest in estimating vaccine effectiveness (VE) in this population following its widespread vaccination. Moreover, because vaccination coverage was so high, it is expected that non-vaccinated persons could be indirectly protected if vaccination reduces infection and transmissibility among vaccinated persons. A few observational studies focusing on the elderly have been published in the last weeks [7,8]; one published and two pre-print studies have specifically addressed vaccine effects in LTCF residents [9,10,11], and none have tried to address the indirect protection in non-vaccinated individuals in this high-coverage setting.

This study aims to estimate indirect and total (direct plus indirect) effects of vaccination in residents in LTCF in a high incidence context.

## Methods

### Data sources

REGVACU is a nation-wide registry of all COVID-19 vaccine-doses administered and vaccine rejections. Data was extracted on March 15 and the administrative censoring date was March 10. Individuals ≥65 years of age by December 27, with a valid postal code, and identified as “resident in elderly homes” according to REGVACU were selected. SERLAB is a nation-wide registry of all SARS-CoV-2 PCR and rapid antigenic tests performed. Positive tests within 60 days of a previous positive one were dropped, as they were considered to belong to the same episode. In LTCF, tests were performed to symptomatic persons and risk contacts. Incoming residents were also routinely tested and periodical screenings have also been carried out. Therefore, documented infections registered in SERLAB may correspond both to symptomatic and asymptomatic infections, although this circumstance was not recorded in the system. Residents in REGVACU were cross-matched with SERLAB by person identification number, date of birth and sex.

### Study design

To estimate the total (direct and indirect) effect of vaccination in vaccinated individuals, the risk of SARS-CoV-2 documented infection in the cohort of individuals with the first dose administered between December 27 and March 10 was compared to the risk in the same individuals in a period before the start of the vaccination programme. A before-after comparison was deemed more appropriate since, due to the high vaccination coverage at LTCFs, non-vaccinated individuals after December 27 would probably not represent baseline infection risk had the individual not been vaccinated. Baseline infection risk, on the other hand, is heavily influenced by community incidence and the vaccination campaign coincided with the third epidemic wave in Spain. To minimize this effect the second epidemic wave was chosen as comparison period, starting the follow-up of the non-vaccinated period 87 days before individual-specific first dose administration date (October 1, at the earliest), with administrative censoring on December 13, 87 days before March 10 (supplementary Figure S1).

To estimate the indirect protection of vaccination in not vaccinated individuals, the risk of SARS-CoV-2 documented infection in the cohort of individuals never vaccinated between December 27 and March 10 was compared to the risk in the same individuals 87 days before, similarly as previously explained for vaccinated individuals. The follow-up period started at the earliest date when the vaccine was offered to each individual, since all residents at the same LTCF were offered vaccination on the same day. Therefore individuals were ensured to be included on the date that a first vaccine-dose was administered to most of the co-residents and workers.

The follow-up for all individuals finished at the earliest of a SARS-CoV-2 positive test or administrative censoring. Unfortunately, no information on the individuals’ vital status was available. Existence of any previous SARS-CoV-2 documented infection on the first day of follow-up was also registered.

An additional analysis to investigate the possible design-associated bias is presented in the supplementary material.

The study obtained approval from the research ethics committee at the Instituto de Salud Carlos III (CEI PI 98_2020). Patients or the public were not involved in the design, or conduct, or reporting, or dissemination plans of our research. Results of this study are planned to be disseminated to the broad public.

### Data analysis

The standardized cumulative risk of a documented SARS-CoV-2 infection that every individual had in the sample been either vaccinated or not vaccinated was computed [12]. To estimate the probability of the event on each follow-up day, conditioned to remaining event-free up to that day and given the individual covariates, a pooled logistic regression was fitted adjusting by follow-up day, previous SARS-CoV-2 infection (before beginning of follow-up), daily-varying 7-day SARS-CoV-2 cumulative incidence specific to the province, its quadratic term, and the empirical reproduction number for that province on that date. An interaction between follow-up day and vaccination was introduced to allow for a time-varying effect of the vaccine. Robust models were built using individuals as clusters. Standardized cumulative risk curves were derived using the Kaplan-Meier method. Risk ratios (RR), vaccine effectiveness (VE= 1-RR) and risk difference (RD) were estimated for the overall period and in four sub-periods after the administration of the first dose, as proxies of different vaccine protection: (1) 14 days; (2) 14 to 21 days; (3) 21 to 28 days (proxy of first 7 days after the second dose) and; (4) >28 days (proxy of fully vaccinated, i.e. ≥7 days after second vaccine dose). Normal distribution-based confidence intervals were estimated using bootstrapping with 300 repetitions.

## Results

### Description of participants

Out of 5,068,733 vaccination records from 3,615,403 individuals in REGVACU, 573,533 records from 299,209 individuals were selected; 296,093 (99.0%) had received ≥1 vaccine-dose, of which 99.8% were Pfizer/BioNTech (BNT162b2) and 0.2% Moderna vaccine; 92.6% of them received a second vaccine-dose in a median of 21 days (interquartile range: 21-21). Time to vaccination is shown in supplementary figure S2. Mean age was 85.9 years (standard deviation = 7.8) and 70.9% were females. Selected individuals were cross-matched with SERLAB; 77,662 (26.0%) had at least one positive test between March 1, 2020 and March 11, 2021. A SARS-CoV-2 previous infection was found in 17.5% of participants on the date they started the follow-up for the total effects study; 22.3% in the vaccinated group and 12.7% in the comparison group (from 87 days before). In the indirect effects analysis, 20.3% had previous infection; 27.7% in the indirectly protected and 12.9% in the comparison group.

### Estimation of vaccine effectiveness in vaccinated persons

This analysis included 16,277,284 and 16,142,536 person-days of follow-up among vaccinated and non-vaccinated persons, respectively. There were 11,304 and 19,656 documented infections, respectively (supplementary table S1). Detailed information on the crude estimates and adjusted cumulative risk in each group can be found in the supplementary material (Figure S3 and Tables S2 and S3).

Vaccine effectiveness for the whole study period was 57.2% (95% Confidence Interval: 56.1% to 58.3%), but it increased after two vaccine doses, and was higher in individuals without previous SARS-CoV-2 infection; VE was 81.8% (81.0% to 82.7%) for residents fully vaccinated and with no previous infection, but decreased to 56.8% (47.1% to 67.7%) if previous infection (Table 1, Figure 1). Interestingly, in a separate analysis we found that previous infection in the reference period was associated to a risk reduction of 86.6% (85.2%-87.8%), higher than the estimate for complete vaccination.

**Table 1.**
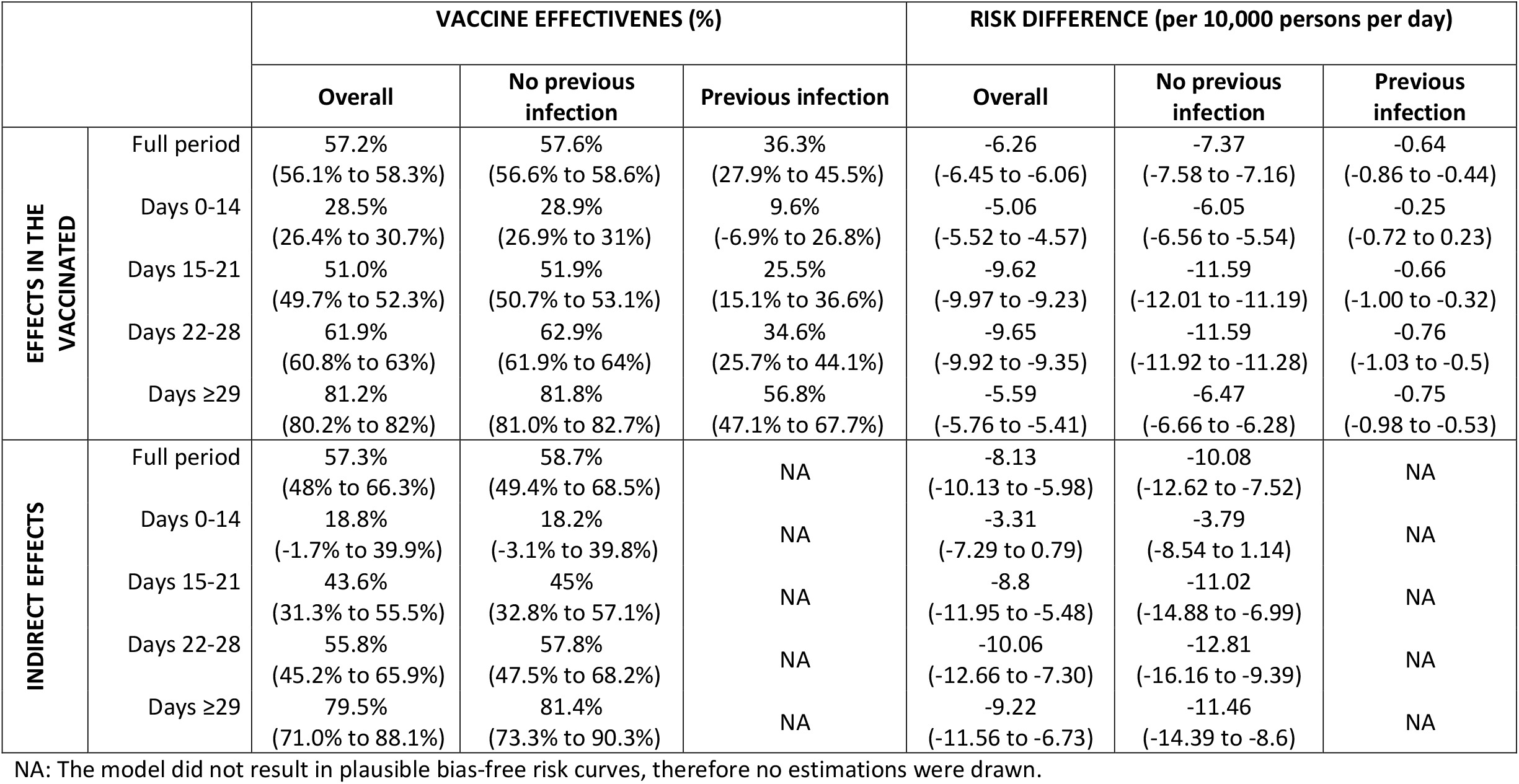
Vaccine effectiveness (VE) and risk difference (RD) in residents of elderly long-term care facilities according to evidence of previous infection and time since first vaccinated (as a proxy of number of vaccine - doses and days since last dose).

**Figure 1.**
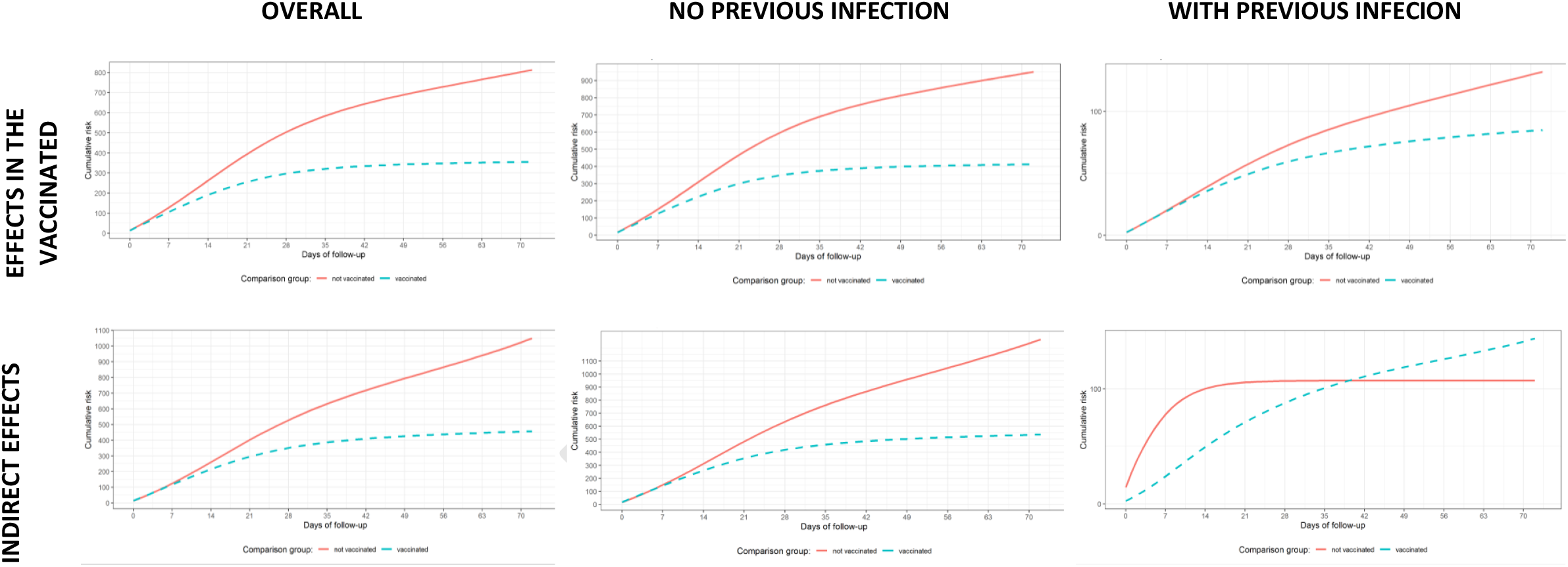
Cumulative incidence of documented SARS-CoV-2 infection in residents in long-term care facilities estimated from adjusted hazards models.

The estimated number of SARS-CoV-2 infections averted by vaccination (risk difference) was greatest in the intermediate periods, which coincided with the peak of the epidemic waves, at 11.6 cases per 10,000 vaccinated persons per day in the group without previous infection (Table 1). In the group with previous infection, the number of infections averted was much lower, of around 0.6 – 0.7 per 10,000 vaccinated persons per day.

### Estimation of indirect vaccine effectiveness in non-vaccinated persons

This analysis included 164,520 and 161,388 person-days of follow-up, respectively, among persons not vaccinated but who had been offered the vaccine at their LTCF (indirectly protected) and same persons in the reference period (87 days before). There were 126 and 276 events, respectively (supplementary Table S1). Detailed information on the crude estimates and the adjusted cumulative risk in each group can be found in the supplementary material (Figure S3 and Tables S2 and S3).

Indirect protection was estimated at 57.3% (48% to 66.3%) for the whole study-period. There was no statistically significant reduction in risk in the first 14 days of follow-up but it increased progressively thereafter, particularly after 28 days (as a proxy of full immunization of vaccinated persons at the LTCF), when VE reached 79.5% (71.0% to 88.1%) overall and 81.4% (73.3% to 90.3%) for the group with no documented SARS-CoV-2 infection before the beginning of follow-up (Table 1, Figure 1).

The estimated number of SARS-CoV-2 infections averted by vaccination was similar to the one found in the vaccinated group for individuals without previous infection, of 11 .0 and 12.8 per 10,000 non-vaccinated persons per day in the intermediate periods (Table 1).

It was not possible to estimate VE for indirect protection in the group with a previous SARS-CoV-2 infection since there were only 14 events, confidence intervals virtually tended to infinite, and the model did not result in credible risk curves.

## Discussion

This study on the institutionalized elderly confirms the high benefit of vaccination in this population, reducing the risk of infection by up to 81.2% and avoiding up to 9.6 cases per 10,000 population per day. The risk reduction was through direct protection of those vaccinated but also through indirect protection of those who were not-vaccinated. The vaccine effectiveness increased throughout the study period, likely showing the progressive immunization of vaccinated persons with increasing time elapsed since the first-dose and after the receipt of the second dose. While VE was higher for individuals naïve to SARS-CoV-2, those with previous infection also benefited from vaccination, even the absolute gain in number of infections averted was low, possibly due to an already lower baseline risk in this group.

Immunesenescence and factors related to chronic conditions, together with malnutrition, are known to impair immunity required for an effective vaccine response [13], and lower neutralizing antibodies response to Pfizer/BioNTech vaccine in people ≥65 years has been reported [6,14]. However, our estimates resulted fairly similar to those of observational studies in younger adult population and are consistent with other studies showing high VE in the elderly from the general population. A cohort of health care workers in the UK found a VE of 70% 21 days after the first dose and of 85% 7 days after the second dose of Pfizer/BioNTech [15]. A slightly higher estimate, of 94.1%, is given by a pre-print with data from Israel [16]. Other observational studies have explored VE in older age groups. In a registries-based study from Israel, in persons aged ≥70 years, VE was found to be 44%, 64% and 98% at 14-20 days post-vaccination, 21-27 days post-vaccination and ≥7 days after the second vaccine-dose, respectively, which were similar to the results for younger age groups [17]. Bernal et al have reported vaccine effects to start 10-13 days after vaccination with Pfizer/BioNTech and reach 61% in people aged ≥70 years and 70% in people aged ≥80 years ≥28 days post-vaccination, and 89% 14 days after the second vaccine-dose [8].

A study in LTCF in Connecticut experiencing COVID-19 outbreaks found a 63% protection with partial vaccination (between 14 and 28 days of the first dose), close to our estimates, with unchanged results after excluding those with previous infection [9]. However, two other existing studies focusing on LTCF have reported lower VE. A Danish study in pre-print [10], has found no protective effect of a first vaccine-dose, a 52% reduction in days 0-7 after the second dose and 64% beyond day 7. A recently released pre-print manuscript from the VIVALDI study in the UK has found no protection conferred by vaccination with the Pfizer/BioNTech vaccine in the first 28 days after the first dose [11]. Nevertheless, VE between days 28 to 47 was between 56% and 62% [11], in a similar range of the effect found in this study for the period 22-28 days (61.9%). Early results from British Columbia have estimated 80% reduction in risk 2-3 weeks after the first vaccine-dose [18]. Of note, our work included both symptomatic and asymptomatic infections, pointing that risk was probably reduced for both type of endpoints to an unknown degree. As an illustration, in national COVID-19 surveillance, 39% of all notified infections since 10 May 2020 in people ≥65 years of age were asymptomatic.

A considerable 22% of all participants in our study had a previous documented SARS-CoV-2 infection, although there are possibly a high number of infections that were not documented, especially during the first epidemic wave in March-April 2020. Several studies have documented a high immune response to a first COVID-19 vaccine-dose in people with previous infection [19,20,21]. The results of this study add to previously existing literature that, even though the effect was greater in naïve subjects to SARS-CoV-2, those with previous infection also benefited from a risk reduction of 57%, although it translated in less than 1 infection averted per 10,000 population per day. Results from the indirect protection analysis support the hypothesis that vaccination may reduce transmissibility of SARS-CoV-2 and result in herd immunity. Previous studies have shown decreased viral load in vaccinated patients, including those in LTCF [9, 22], and a study from Scotland found a 30% lower risk of SARS-CoV-2 in household members of vaccinated health-care workers, although the reduction in SARS-CoV-2 transmission from vaccinated individuals could be double that estimate, since household members could also have been infected in the community [23]. A recent ecological study from Israel has shown that increasing vaccine coverage provides cross-protection to unvaccinated individuals in the community [24]. In our study, non-vaccinated individuals living in facilities where the majority (residents and staff) had been vaccinated showed a risk reduction similar to those actually vaccinated. However, the magnitude of protection may be overestimated, since non-vaccinated individuals could correspond more frequently to persons with previous infection, even if not documented. This could be controlled in individuals with documented infection but not in an unknown number with non-documented infection or diagnosed with serology. Also, indirect protection was measured in a context of very high vaccine coverage, difficult to attain in a non-institutional setting; therefore our results may not have generalisability to the community setting.

Some limitations to study results could relate to the before-after comparison. Even though we tried to minimize it, residual confounding due to higher incidence during the third epidemic wave and possibly, to the relaxation of the isolation of LTCF during the Christmas season, with higher number of day-outs and visits, may be present and could underestimate the protection of the vaccine. This underestimation of the effect of the vaccine could maybe explain that the effect of natural infection in the non-vaccinated group was found higher than the effect of the vaccine. The conservative direction of the possible bias is shown by the bias indicator analysis (supplementary material).

In conclusion, our results confirm the effectiveness of vaccination in institutionalized elderly population, endorse the policy of universal vaccination in this setting, including in people with previous infection, and suggest that even non-vaccinated individuals benefit from indirect protection. Further questions include the duration of protection in this population and according to previous infection, and the severity of infection, which could not be measured in this study.

## Data Availability

Authors could make study data available upon request

## Acknowledgements

We would like to acknowledge the contribution of the General Directorate of Digital Health and National Health Service Information Systems and the General Sub-Directorate for Health Information (Ministry of Health, Spain) for their work in managing the registries in which this study was based, as well as in providing all the necessary IT support. We specially thank Miguel Hernán for his methodological advice and support in the design of the analysis as well as Luis Sanguiao for his support in implementing parallel computing. We acknowledge as well all the work of the professionals involved in the vaccination program and in managing the vaccination and laboratory registries from the Autonomous Communities.

