## Supplementary information for "Direct and indirect effectiveness of mRNA vaccination against SARS-CoV-2 infection in long-term care facilities in Spain"

**Figure S1. Seven-days cumulative incidence of SARS-CoV-2 newly diagnosed patients in Spain, and in shadowed areas, the study period for the selected individuals (December 27, 2020 – March 10, 2021) and the reference period 87 days before(October 1 to December 13, 2020).**

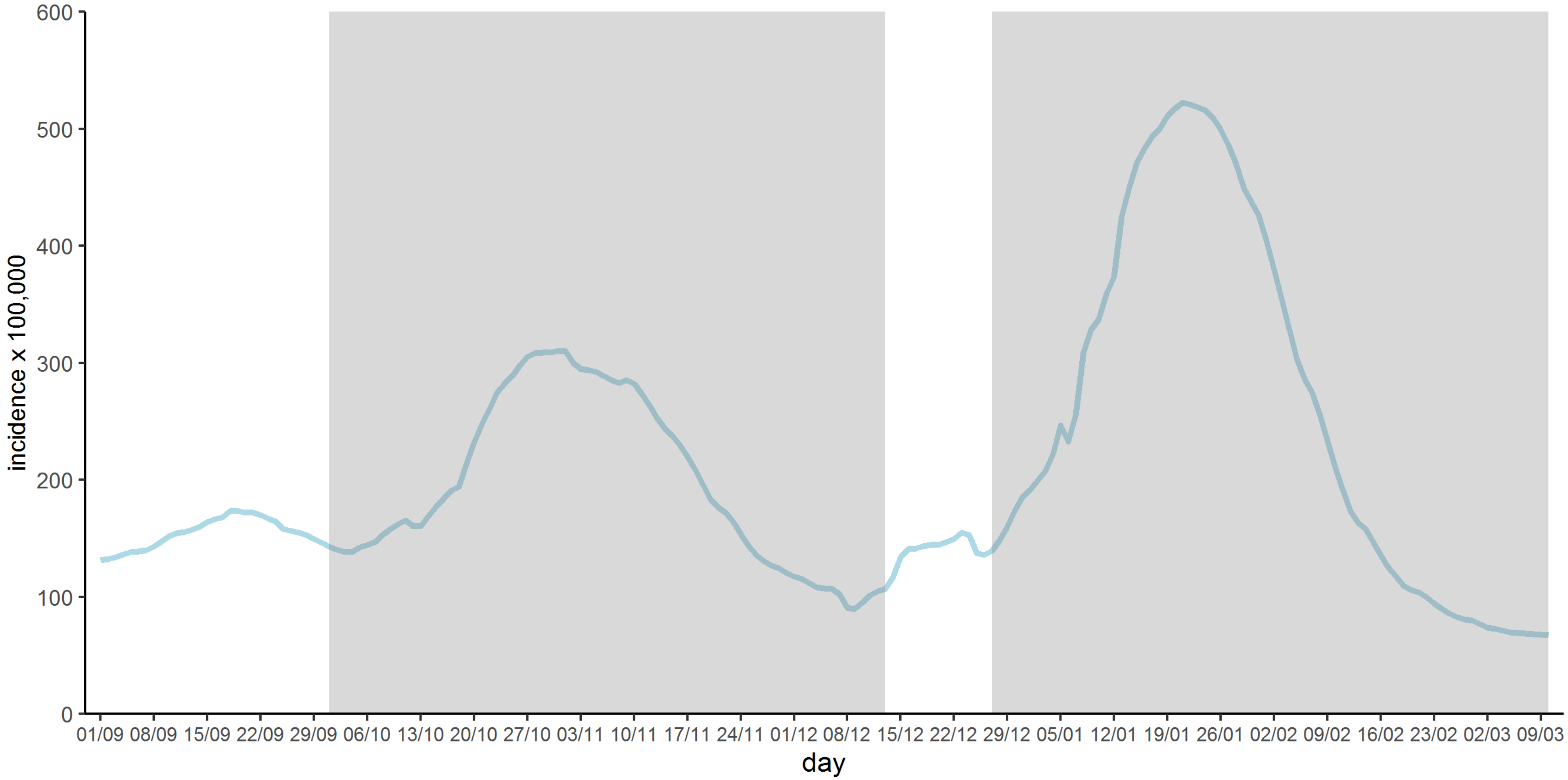

**Figure S2. Vaccination uptake in residents in long-term care facilities included in the study (N=299,209), starting on 27 December, 2021.**

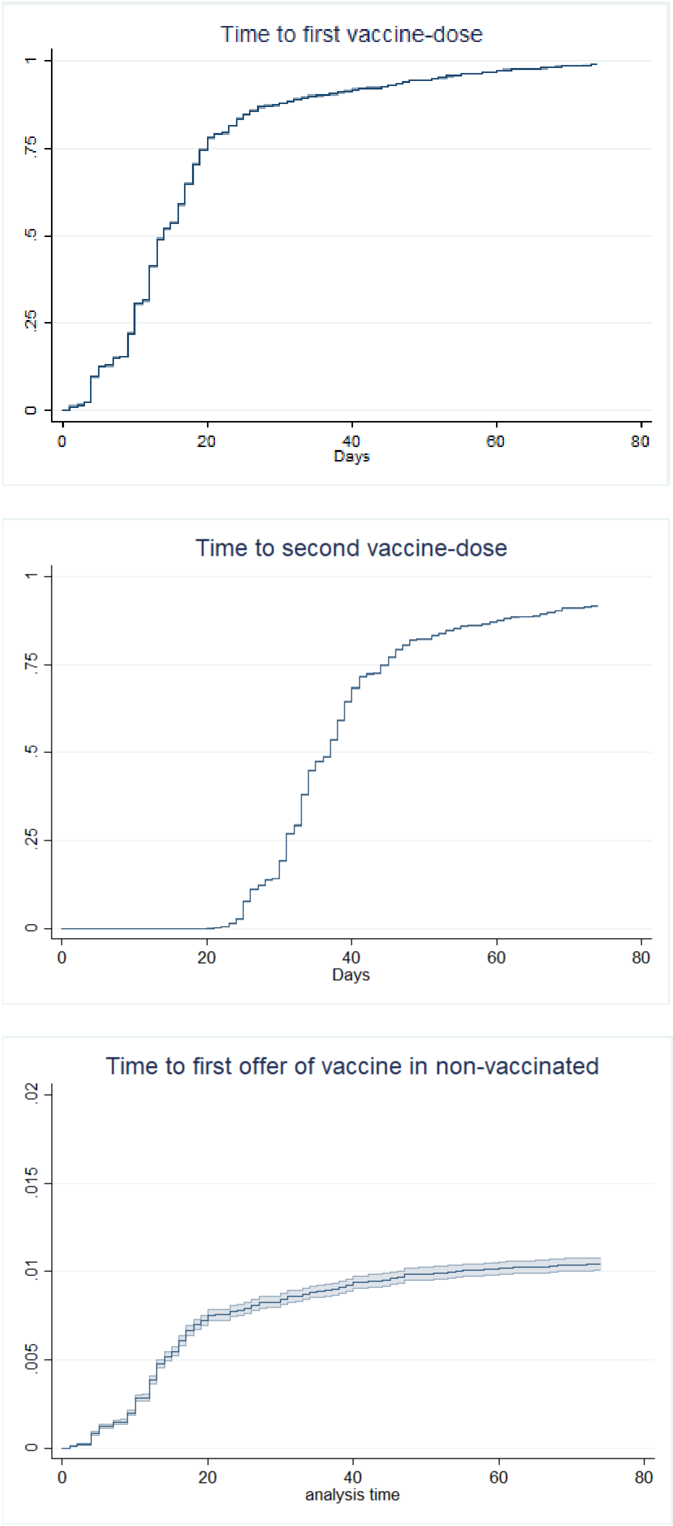

**Figure S3. Unadjusted Kaplan-Meier estimates of the cumulative risk of a positive SARS-CoV-2 diagnosis in vaccinated individuals vs. non-vaccinated individuals from the reference period (left) and in non-vaccinated patients after being offered vaccination (indirectly protected) vs. non-vaccinated individuals from the reference period (right).**

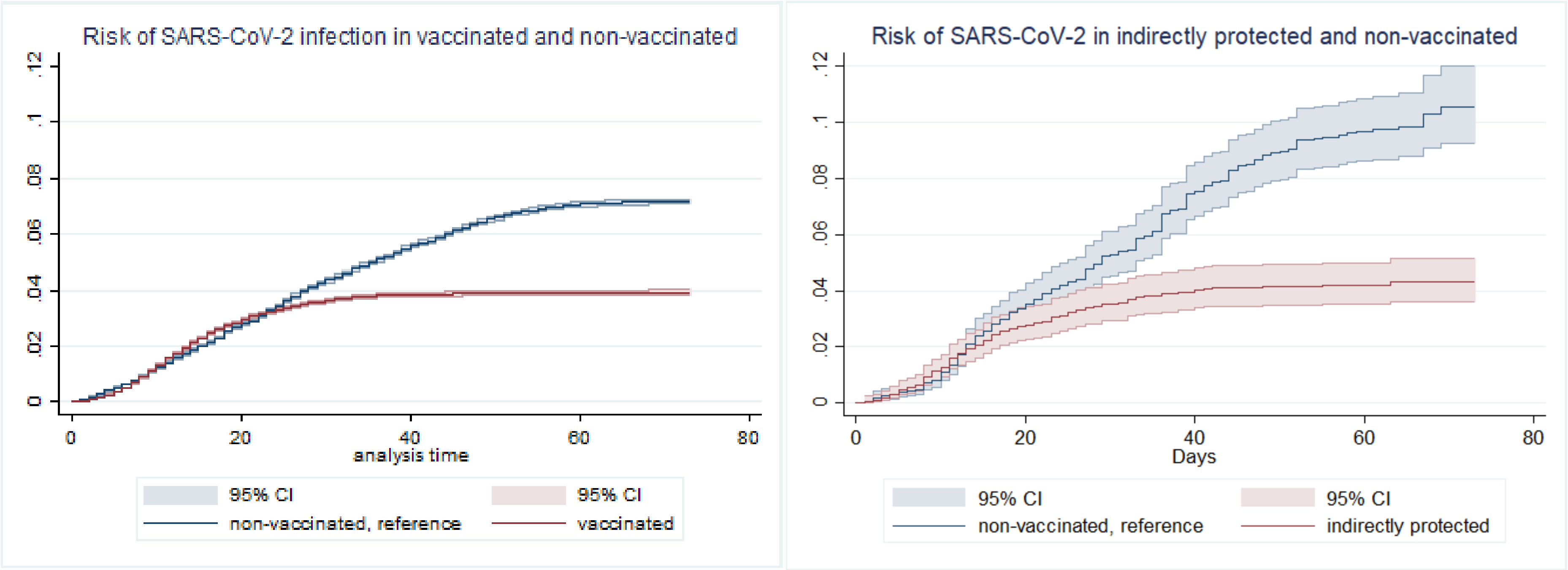

**Table S1. Person-days of follow-up and number of events in the different groups and study periods.**

| | | Person-days | | Events | |
|---|---|---|---|---|---|
| | | No Vac* | Vac* | No Vac* | Vac* |
| EFFECTS IN THE VACCINATED | Full period | 16,142,536 | 16,277,284 | 19,656 | 11,304 |
| | Days 0-14 | 4,076,665 | 4,076,391 | 6,202 | 5,455 |
| | Days 15-21 | 1,963,952 | 1,969,388 | 2,794 | 3,038 |
| | Days 22-28 | 1,910,859 | 1,908,664 | 1,308 | 3,311 |
| | Days 29-73 | 8,325,808 | 8,188,093 | 1,000 | 7,852 |
| INDIRECT EFFECTS | Full period | 161,388 | 164,520 | 276 | 126 |
| | Days 0-14 | 42,854 | 42,820 | 74 | 63 |
| | Days 15-21 | 20,450 | 20,552 | 38 | 24 |
| | Days 22-28 | 19,746 | 19,966 | 38 | 17 |
| | Days 29-73 | 78,338 | 81,182 | 126 | 22 |

*Vac: For the Effects in the Vaccinated it corresponds to vaccinated persons; for the Indirect effect it corresponds to non-vaccinated persons after vaccine introduction in their long-term care facility; No Vac: corresponds to the non-vaccinated reference period (87 days before first vaccination or before offering of the vaccine).

**Table S2. Estimated standardized cumulative risk (per 10,000 persons per day) and 95% confidence interval in the different groups and study periods, from adjusted hazard models.**

| | | Vaccinated / Indirectly protected (study group) | Not vaccinated (reference) |
|---|---|---|---|
| EFFECTS IN THE VACCINATED | Full period | 4.68<br>(4.6 to 4.77) | 10.94<br>(10.79 to 11.09) |
| | Days 0-14 | 12.66<br>(12.42 to 12.89) | 17.72<br>(17.32 to 18.09) |
| | Days 15-21 | 9.24<br>(9.06 to 9.42) | 18.85<br>(18.53 to 19.15) |
| | Days 22-28 | 5.93<br>(5.79 to 6.08) | 15.58<br>(15.33 to 15.82) |
| | Days 29-73 | 1.3<br>(1.25 to 1.35) | 6.88<br>(6.72 to 7.05) |
| INDIRECT EFFECTS | Full period | 6.06<br>(5.02 to 7.15) | 14.19<br>(12.3 to 15.98) |
| | Days 0-14 | 14.31<br>(11.48 to 17.16) | 17.62<br>(14.49 to 20.65) |
| | Days 15-21 | 11.39<br>(9.49 to 13.5) | 20.19<br>(17.52 to 22.9) |
| | Days 22-28 | 7.96<br>(6.36 to 9.71) | 18.02<br>(15.69 to 20.34) |
| | Days 29-73 | 2.38<br>(1.49 to 3.27) | 11.6<br>(9.19 to 13.85) |

* Non-vaccinated persons after vaccine introduction in their long-term care facilit

**Table S3. Crude estimations: Vaccine effectiveness (VE) and risk difference (RD) in residents of elderly long-term care facilities according to evidence of previous infection and time since first vaccinated (as a proxy of number of vaccine - doses and days since last dose).**

| | | VACCINE EFFECTIVENES (%) | | | RISK DIFFERENCE (per 10,000 persons per day) | | |
|---|---|---|---|---|---|---|---|
| | | Overall | No previous infection | Previous infection | Overall | No previous infection | Previous infection |
| EFFECTS IN THE VACCINATED | Full period | 50.8% (49.6% to 52%) | 46,5% (45,3% to 47,7%) | 34,1% (25,6% to 43,4%) | -5.33 (-5.51 to -5.14) | -5,48 (-5,68 to -5,28) | -0,59 (-0,8 to -0,39) |
| | Days 0-14 | 3.4% (0.7% to 5.9%) | -6,3% (-8,9% to -3,6%) | -6,4% (-24,8% to 12,9%) | -0.53 (-0.94 to -0.11) | 1,13 (0,66 to 1,58) | 0,15 (-0,28 to 0,59) |
| | Days 15-21 | 39.7% (38.2% to 41.2%) | 34,8% (33,3% to 36,2%) | 18,4% (7,3% to 30,2%) | -5.31 (-5.56 to -5.05) | -5,22 (-5,49 to -4,96) | -0,39 (-0,67 to -0,11) |
| | Days 22-28 | 55.3% (54.1% to 56.5%) | 52,1% (50,9% to 53,3%) | 31,3% (22,2% to 41%) | -6.65 (-6.85 to -6.43) | -7,03 (-7,26 to -6,8) | -0,61 (-0,85 to -0,38) |
| | Days ≥29 | 81.1% (80.3% to 81.8%) | 80,1% (79,3% to 80,9%) | 59,7% (51,3% to 69,1%) | -6.62 (-6.81 to -6.43) | -7,33 (-7,53 to -7,12) | -0,85 (-1,09 to -0,63) |
| INDIRECT EFFECTS | Full period | 62.3% (53.9% to 70.4%) | 57.9% (48.3% to 67.8%) | NA | -9.47 (-11.65 to -7.14) | -10.03 (-12.65 to -7.4) | NA |
| | Days 0-14 | 22.1% (2.7% to 42.2%) | 7.8% (-15.2% to 31.7%) | NA | -4.14 (-8.27 to 0.06) | -1.62 (-6.52 to 3.37) | NA |
| | Days 15-21 | 48.6% (37.2% to 59.7%) | 41.4% (28.5% to 54.5%) | NA | -8.44 (-11.18 to -5.56) | -8.05 (-11.19 to -4.83) | NA |
| | Days 22-28 | 60.6% (51.2% to 69.6%) | 56.1% (45.7% to 66.8%) | NA | -10.01 (-12.4 to -7.49) | -10.46 (-13.23 to -7.67) | NA |
| | Days ≥29 | 82.6% (76% to 89.1%) | 81.8% (74.8% to 89.5%) | NA | -11.2 (-13.71 to -8.52) | -12.89 (-15.88 to -9.91) | NA |

### Bias indicator analysis

To check the possible design-associated bias, we created a bias-indicator cohort with non-vaccinated time-at-risk of individuals who were later vaccinated, with follow-up starting on December 27 and ending at date of first vaccine dose or date of SARS-CoV-2 infection. We compared it to equivalent follow-up periods of the non-vaccinated cohort from 87 days before, in the same way as exposed in the main text.

Crude risk of infection resulted much higher in bias-indicator cohort (Figure S4), with an overall estimated effect of 1.71 (95%CI: 1.62 – 1.81). This bias was mitigated but not eliminated after adjusting by the same variables as in the main analysis, with an adjusted estimator of 1.36 (95%CI: 1.27 – 1.46). This shows the high incidence in LTCF during the beginning of the third wave, in a moment when many LTCF had not yet been reached by vaccination. In fact, the proportion of persons with past COVID-19 infection increased from around 13% in the reference period to around 25% on the individuals recruited after December 27, showing the high incidence in these facilities. As discussed in the main text, these results indicate the possibility of a residual bias that nevertheless would be conservative, in the sense that it would mean a higher risk in the period when individuals got vaccinated.

**Figure S4. Unadjusted Kaplan-Meier estimates of the cumulative risk of a positive SARS-CoV-2 diagnosis in the bias indicator cohort (left) and adjusted estimates of cumulative risk (right)**

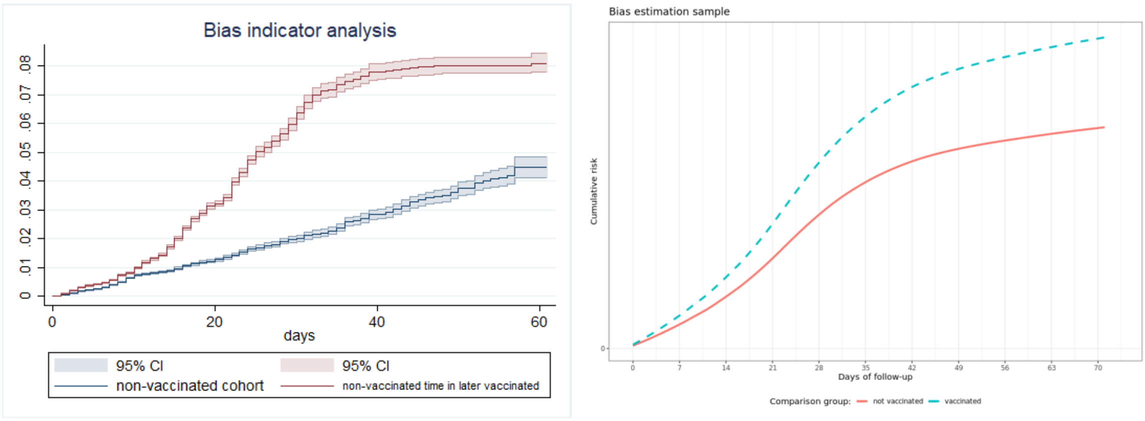
